# Could the Inflation Reduction Act Maximum Fair Price Hurt Patients?

**DOI:** 10.1101/2024.06.26.24309544

**Authors:** Esteban Rivera, Anne M. Sydor, Robert Popovian

## Abstract

**Background:** The Inflation Reduction Act’s Medicare Drug Price Negotiation Program allows the federal government to negotiate caps for select medications. These price caps may reduce revenue for the pharmacy benefit managers (PBMs) that negotiate the actual price paid for medicines in the US. To offset the resulting pressure on their profit margins, it is possible that PBMs would, in turn, increase patient’s out-of-pocket costs for medicines with capped prices. The model presented here evaluates how such increased out-of-pocket costs for the anticoagulants Eliquis (apixaban) and Xarelto (rivaroxaban) could impact patients financially and clinically.

**Methods:** Copay distributions for all 2023 prescription fills for Eliquis and Xarelto managed by the three largest PBMs were used to approximate current copay costs. Increased out-of-pocket costs were modeled as a shift of all Eliquis and Xarelto prescriptions to the highest copay tier. The known linear relationship between copay costs and treatment abandonment was used to calculate the potential resulting increase in treatment abandonment. Known rates of morbidity and mortality due to abandoning anticoagulants were used to estimate resulting increases in morbidity and mortality.

**Results:** If the three largest PBMs all shifted costs onto patients by moving all Eliquis and Xarelto prescriptions to the highest formulary tier, Tier 6, patients’ copay amount would increase by $235 to $482 million for Eliquis and $105 to $206 million for Xarelto. Such an increase could lead to 169,000 to 228,000 patients abandoning Eliquis and 71,000 to 93,000 abandoning Xarelto. The resulting morbidity and mortality could include up to an additional 145,000 major cardiovascular events and up to 97,000 more deaths.

**Conclusion:** The Medicare Price Negotiation Program could impact patients negatively if it causes PBMs to increase patients’ out-of-pocket costs for medicines. Policymakers should closely monitor changes in overall affordability, including all patient out-of-pocket expenditures, for medications in the program. Preemptive measures to ensure that the most vulnerable citizens are not placed in precarious situations leading to poorer health outcomes should be considered.

## INTRODUCTION

As with any complex U.S. federal legislation, the Inflation Reduction Act (IRA), signed into law on August 16, 2022, includes provisions with both positive and negative consequences for diverse stakeholders.^1^ Specific IRA clauses affect health insurance premiums, out-of-pocket healthcare costs for patients, and wholesale medication prices. These changes will impact patients, caregivers, healthcare professionals, biopharmaceutical companies, pharmacy benefit managers (PBMs), health insurance industries, and policymakers differently, depending on their perspectives and roles within the healthcare system, leading to varying positive and negative effects across these groups.

Eliminating out-of-pocket vaccination costs for seniors is an example with mostly positive benefits for all affected groups for whom costs will be reduced, and access to potentially life-saving interventions will be increased.^2^ A shift to preventive care, rather than sick care, will benefit healthcare systems and providers and substantially reduce healthcare costs.^3^ Conversely, other policy shifts, such as the requirement to cap out-of-pocket costs at $2,000 and negotiate wholesale price limits for seniors may not be as beneficial as propagated. For example, limiting out-of-pocket expenses will benefit the 20% of Medicare enrollees who currently pay more than that.^4^ However, this policy change may also incentivize PBMs to increase costs to that upper limit for patients who currently pay less than that threshold. In addition, PBMs may also increase burdensome utilization management techniques to offset their increased costs.^5–7^

### Medicare Drug Price Negotiation Program

The IRA authorized the Secretary of the Department of Health and Human Services (HHS) to negotiate a “maximum fair price (MFP)” directly with manufacturers for some of the most highly used brand-name medications for patients paid for by Medicare.^1^ Many parties have raised concerns about this price setting.^8–11^ An analysis from the Congressional Budget Office (CBO) concluded that drug price setting would limit innovation, stall research and development, and likely reduce the number of new drug applications to the FDA. ^12^

At face value, lower wholesale medication prices appear purely beneficial to patients and harmful to biopharmaceutical manufacturers. However, this does not account for how medicines are purchased in the U.S. healthcare system. In reality, the wholesale list price is rarely, if ever, the amount that a biopharmaceutical company receives for a medication. Almost all prescription medications are purchased under an agreement negotiated by a PBM. These agreements grant substantial concessions, such as fees, discounts, and rebates, to the health insurer and PBMs.^13^ Notably, the net price paid for medicines is not publicly available, and it is impossible to know precisely how a wholesale price reduction will affect the actual cost of medications for patients.^14^ It should also be noted that the negotiated discounts, rebates, and fees are typically not passed on to patients.^15, 16^

Negotiations between PBMs and manufacturers regarding the fees, rebates, and discounts manufacturers will give PBMs are based on the manufacturer’s wholesale list price. Higher list prices create more room for negotiating concessions, whereas lowered, set prices will likely result in reduced concessions from manufacturers to the PBMs.^17^ Such dynamics increase financial pressure on PBMs and insurers (often owned by the same parent company). In addition, IRA also caps out-of-pocket costs, increases plan liabilities, and restricts Medicare Part D premium increases, thus curtailing the ability of PBMs to seek profits elsewhere.^1^ In short, the Medicare Drug Price Negotiation Program may negatively affect PBM profits. Ultimately, patients may be negatively impacted as the PBMs will find new schemes to make up lost revenue and profits by shifting greater costs to the patients or restricting access to life-saving medicines.

Over the past few decades, PBMs have increased their management of access to medications with tiered formularies, which exchange coverage of specific treatments for higher rebates and fees from biopharmaceutical manufacturers.^18^ Under the current rebate contracting methodology, PBMs are incentivized to cover higher-priced treatments because these result in more significant rebates and fees for the PBM. This misalignment of incentives has led to the exclusion of hundreds of medicines from PBM formularies or the placement of medications that do not benefit the bottom line of PBMs on higher tiers where patient out-of-pocket costs are more significant. The practice of formulary exclusions and increasing out-of-pocket expenses by moving medications to a formulary tier that requires patients to pay more out of pocket can harm both patients’ pocketbooks and health outcomes.^19, 20^ Such tactics by the PBMs have led to patients abandoning their treatments for nonmedical reasons (e.g., financial burdens), leading to adverse events and increased mortality and morbidity. ^21–23^ The number of patients impacted is not inconsequential. Published studies have demonstrated that specific formulary exclusions and changes in out-of-pocket requirements impact hundreds of thousands of patients.^19–23^ In addition, a study by IQVIA illustrates a linear correlation between the copay costs of medication and the proportion of patients who abandon it.^24^

## PURPOSE OF THIS STUDY

This study aims to achieve three objectives by analyzing the impact of price setting for two anticoagulants, Eliquis and Xarelto, which are among the first ten drugs selected for the Medicare Drug Price Negotiation Program.

1. Estimate the potential increase in out-of-pocket costs for Medicare Part D enrollees if the three major PBMs, CVS Caremark (CVS), Optum RX (ORx), and Express Scripts International (ESI)—which together negotiate actual prices for 80% of all prescriptions in the US^25^—increase patients’ out-of-pocket costs for Eliquis or Xarelto.
2. Estimate the number of Medicare Part D enrollees who may abandon their treatments with Eliquis or Xarelto due to higher out-of-pocket costs.
3. Estimate the potential number of Medicare Part D enrollees who will suffer consequential adverse health outcomes because of abandoning their treatment with Eliquis or Xarelto.

Understanding these effects is crucial for policymakers to ensure that the IRA achieves its cost-saving goals without compromising patient access to essential treatments.

## METHODS

We evaluated the potential economic impact if CVS, ORx, and ESI, the three largest PBMs, increase out-of-pocket requirements for Eliquis and Xarelto. We utilized switching tiers as a surrogate to estimate how and by how much patients’ out-of-pocket requirements could change. We modeled how moving Eliquis or Xarelto to Tier 6 from Tier 3 in the ESI, CVS, or ORx formularies would affect: 1) the increased dollar amounts patients would pay for these medications annually; 2) the number of people who would likely stop taking their prescribed Eliquis or Xarelto; and 3) the potential increases in morbidity or all-cause mortality due to increased abandonment rates.

### Data Sources, Structure, and Key Variables

Rates of abandonment, morbidity, and mortality were based on published studies.^21–23^ IQVIA data was utilized in estimating out-of-pocket expenditures and abandonment (discontinuation of prescriptions) correlations.^24^ Pfizer Inc. supplied unpublished data on the number of total Medicare Part D prescriptions by formulary tier and copay cost ranges in 2023 for all Eliquis and Xarelto prescription fills managed by the three largest PBMs (CVS, ORx, and ESI).

### Model Assumptions

A switch to a higher tier was used as a surrogate to develop a model estimating out-of-pocket cost changes, assuming all Tier 3 prescriptions would become Tier 6. Rather than theorizing what PBM behavior might be, we used prior PBM behavior (copay range structure in 2023) as a proxy for increased out-of-pocket costs. Because only ESI had a Tier 6 copay structure, it was also assumed that behavior among the three PBMs would be similar. When modeling an increase in out-of-pocket costs for CVS and ORx Tier 3 prescriptions, the Tier 6 copay structure for ESI was used. The midpoint of a Tier copay range was used to calculate total costs. For example, a copay of $75 was used for the $50-$100 range. All out-of-pocket cost increases were dependent on the difference in structure between Tier 3 and Tier 6 by ESI in 2023. To model potential abandonment and increases in morbidity and mortality, we assumed a 90- day prescription refill across all data and thus divided total annual prescriptions (Pfizer unpublished data) by four to estimate the total number of people who took either Eliquis or Xarelto in 2023. Rates of abandonment after copay increases from IQVIA data were used to estimate how many people would abandon treatment with Eliquis or Xarelto within each out-of-pocket dollar amount.^24^ A 45% and 30% increase, respectively, in the likelihood of major cardiovascular events and all-cause mortality in people who abandon anticoagulant treatment from a study by Rivera-Caravaca et al.^21^ was used to estimate resulting increases in morbidity and mortality.

### Modeling

Total out-of-pocket costs were dependent on 2023 total prescription fills (TRx), the percentage of total prescriptions within a given tier (%TRx), and the percentage breakdown of each copay range within each tier (%TTRx). Total prescription costs for each Tier were calculated as the sum of the cost per range across all five copay tiers, with cost per range calculated as:

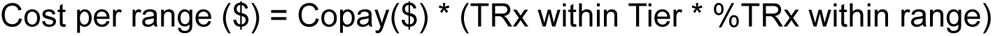

Current costs were calculated based on actual copay structures for each PBM in 2023. Potential costs were calculated, assuming all Tier 3 prescriptions would be switched to Tier 6, using the ESI Tier 6 copay structure. The difference in the copay range structure of Tier 3 and Tier 6 determined the cost differences per range. The total change in out-of-pocket costs was calculated as the difference between the potential costs if Eliquis and Xarelto were moved to formulary Tier 6 and the 2023 copay costs as calculated above.

We calculated the patient abandonment rate of treatment per copay range. The abandonment rate per copay range was calculated to estimate total abandonment. Using the relationship between out-of-pocket-costs and abandonment from IQVIA,^24^ a simple linear regression was estimated as:

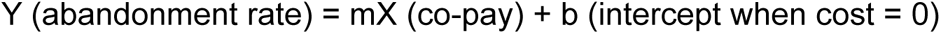

Total prescriptions per range were divided by four because assuming prescriptions were for 90-day refills, every four prescriptions is equivalent to one person using Eliquis or Xarelto during 2023. The calculated coefficient was used to estimate the abandonment rate for each copay range. Rates were estimated based on the midpoint and maximum of each copay range to approximate the number of people likely to abandon treatment because of increased out-of-pocket costs. These rate estimates were multiplied by total prescriptions within each copay range to calculate total abandonment for each range as:

Total abandonment per range = Predicted abandonment * TRx within copay range The increase in the number of people who would abandon Eliquis or Xarelto treatment was calculated as the difference in the total abandonments if all prescriptions were moved to Tier 6 and the likely abandonment rate in 2023. Increased morbidity and mortality were calculated as 45% and 30%, respectively, of the increased abandonments likely to occur if all Eliquis and Xarelto prescriptions were moved to Tier 6.^21^

## RESULTS

### Potential Cost Increases

The total number of Medicare Part D prescriptions for Eliquis and Xarelto in Tier 3 of the CVS, ORx, and ESI formularies are shown in Table 1.

**Table 1.** The number of prescriptions for Eliquis and Xarelto managed by the three largest PBMs as formulary Tier 3 and Tier 6 in 2023.

|  |  |  |  |  |
| --- | --- | --- | --- | --- |
|  | **CVS Tier 3** | **ORx Tier 3** | **ESI Tier 3** | **ESI Tier 6<sup>a</sup>** |
| **Eliquis** | 8,125,970 | 5,895,443 | 3,463,353 | 126,929 |
| **Xarelto** | 2,968,237 | 1,985,963 | 1,195,899 | 43,829 |
| <sup>a</sup>Only ESI currently uses Tier 6 for Eliquis and Xarelto prescriptions. Abbreviations: CVS Caremark, CVS; Express Scripts International, ESI; ORx, Optum Rx; PBM, pharmacy benefit manager. |  |  |  |  |

The copay that patients must cover out of pocket varies by the details of their prescription benefits plan and is only available for ranges of copay values, as shown in Table 2. Notably, differences in the proportion of patients with different copay ranges range from 0.5% to 24.2%.

**Table 2.** The proportion (%) of prescriptions for Eliquis and Xarelto by tier and copay range in the three largest PBMs.

|  |  |  |  |  |  |
| --- | --- | --- | --- | --- | --- |
| **Drug/Co-pay range** |  | **CVS Tier 3** | **ORx Tier 3** | **ESI Tier 3** | **ESI Tier 6<sup>a</sup>** |
| **Eliquis** | \$0 | 25.3 | 27.3 | 24.8 | 18.3 |
| | >\$0-50 | 29.4 | 26.3 | 30.7 | 30.7 |
| | \$50-100 | 25.9 | 30.6 | 6.6 | 8.4 |
| | \$100-200 | 9.9 | 9.3 | 27.6 | 33.5 |
| | >\$200 | 9.5 | 6.5 | 10.3 | 9.0 |
| **Xarelto** | \$0 | 23.4 | 28.6 | 26.6 | 17.0 |
| | >\$0-50 | 25.4 | 22.4 | 28.3 | 27.6 |
| | \$50-100 | 26.3 | 29.7 | 6.3 | 8.7 |
| | \$100-200 | 13.8 | 12.2 | 26.8 | 33.4 |
| | >\$200 | 11.1 | 7.1 | 12.0 | 13.3 |
| <sup>a</sup>Only ESI currently uses Tier 6 for Eliquis and Xarelto prescriptions. Abbreviations: CVS Caremark, CVS; Express Scripts International, ESI; ORx, Optum Rx; PBM, pharmacy benefit manager. |  |  |  |  |  |

Because only ESI uses Tier 6 for Medicare Part D Eliquis and Xarelto prescriptions presently, we used this copay distribution when projecting how out-of-pocket costs could change by all three of the largest PBMs (Table 3).

**Table 3.** Calculated copay costs (2023 $USD) for Medicare Part D Eliquis and Xarelto prescriptions managed by the three largest PBMs.

|  |  |  |  |  |  |  |  |
| --- | --- | --- | --- | --- | --- | --- | --- |
| **Drug** | **PBM** | **Calculated copay costs (2023 \$USD millions)** | | | | | |
|  |  | **2023** |  |  | **If all moved to Tier 6** |  |  |
|  |  | Minimum | Midpoint | Maximum | Minimum | Midpoint | Maximum |
| Eliquis | CVS | 347 | 532 | 723 | 460 | 705 | 958 |
|  | ORx | 225 | 351 | 482 | 333 | 512 | 695 |
|  | ESI | 181 | 276 | 374 | 195 | 301 | 408 |
|  | **Top Three PBMs** | **753** | **1159** | **1579** | **988** | **1518** | **2061** |
| Xarelto | CVS | 148 | 221 | 296 | 194 | 287 | 384 |
|  | ORx | 83 | 127 | 172 | 130 | 192 | 257 |
|  | ESI | 66 | 98 | 132 | 78 | 116 | 155 |
|  | **Top Three PBMs** | **297** | **446** | **590** | **402** | **595** | **796** |
| Abbreviations: CVS Caremark, CVS; Express Scripts International, ESI; ORx, Optum Rx; PBM, pharmacy benefit manager. |  |  |  |  |  |  |  |

Using the cost-range midpoints, the cumulative copay increase from all three of the largest PBMs shifting Medicare Part D prescriptions to Tier 6 (Supplemental Table 1) would be $359 million (CVS: $173M, ORx: $160M, and ESI: $25M) for Eliquis and $149 million (CVS: $66M, ORx $65M, and ESI: $18M) for Xarelto. The differing amounts between the PBMs reflect differences in the number of prescriptions filled by each PBM (Table 1) and the varied distributions of patients within copay ranges for each PBM (Table 2).

**Supplemental Table 1.** Differences in midpoint copay costs if Eliquis and Xarelto moved to Tier 6 (2023 $USD millions).

|  |  |  |  |  |
| --- | --- | --- | --- | --- |
| **Drug** | **PBM** | **Calculated copay costs (2023 \$USD millions)** | | **Difference (all Tier 6 vs 2023 actual)** |
|  |  | **2023** | **All Tier 6** |  |
| Eliquis | CVS | 532 | 705 | 173 |
|  | ORx | 351 | 512 | 161 |
|  | ESI | 276 | 301 | 25 |
|  | **Top Three PBMs** | **1159** | **1518** | **359** |
| Xarelto | CVS | 221 | 287 | 66 |
|  | ORx | 127 | 192 | 65 |
|  | ESI | 98 | 116 | 18 |
|  | **Top Three PBMs** | **446** | **595** | **149** |

Abbreviations: CVS Caremark, CVS; Express Scripts International, ESI; ORx, Optum Rx; PBM, pharmacy benefit manager.

### Abandonment, Mortality, and Morbidity

Data from IQVIA shows a linear relationship between copay amounts and the proportion of patients who stop using a medication,^24^ which we used to calculate the best linear fit slope and coefficient (Figure 1).

**Figure 1.**
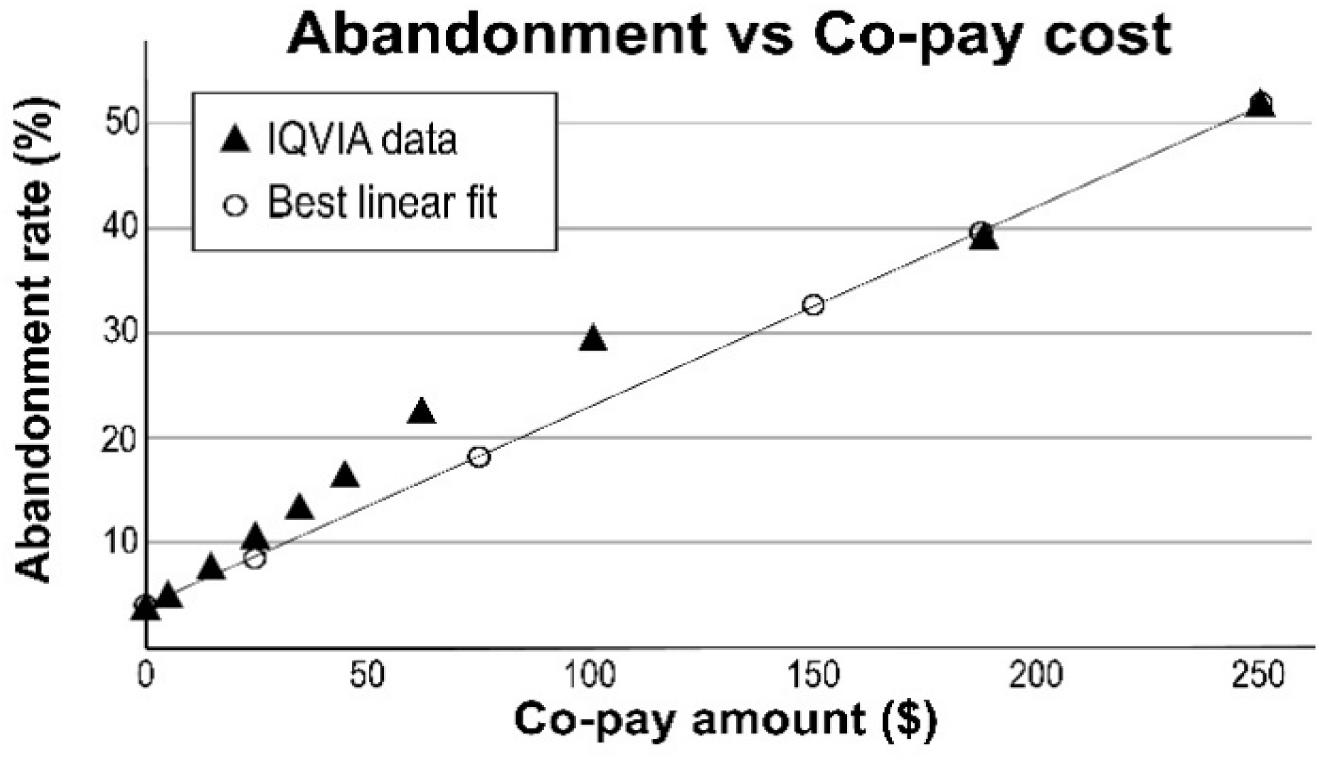
As copay costs rise, the proportion of patients who abandon their treatments with prescription medications rises in a dependent, linear fashion (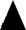).^24^ The best linear fit (◯) was used to calculate the slope and coefficient so those values could be used to predict changes in the abandonment rate for Eliquis or Xarelto if those prescriptions were moved to Tier 6.

After adjustment of prescriptions to approximate the number of people taking Eliquis or Xarelto in 2023 (see Methods), we used the slope and coefficient to estimate the increase in people likely to abandon treatment if Eliquis or Xarelto were shifted to higher out-of-pocket requirement by the CVS, ESI, and ORx (Figure 2).

**Figure 2.**
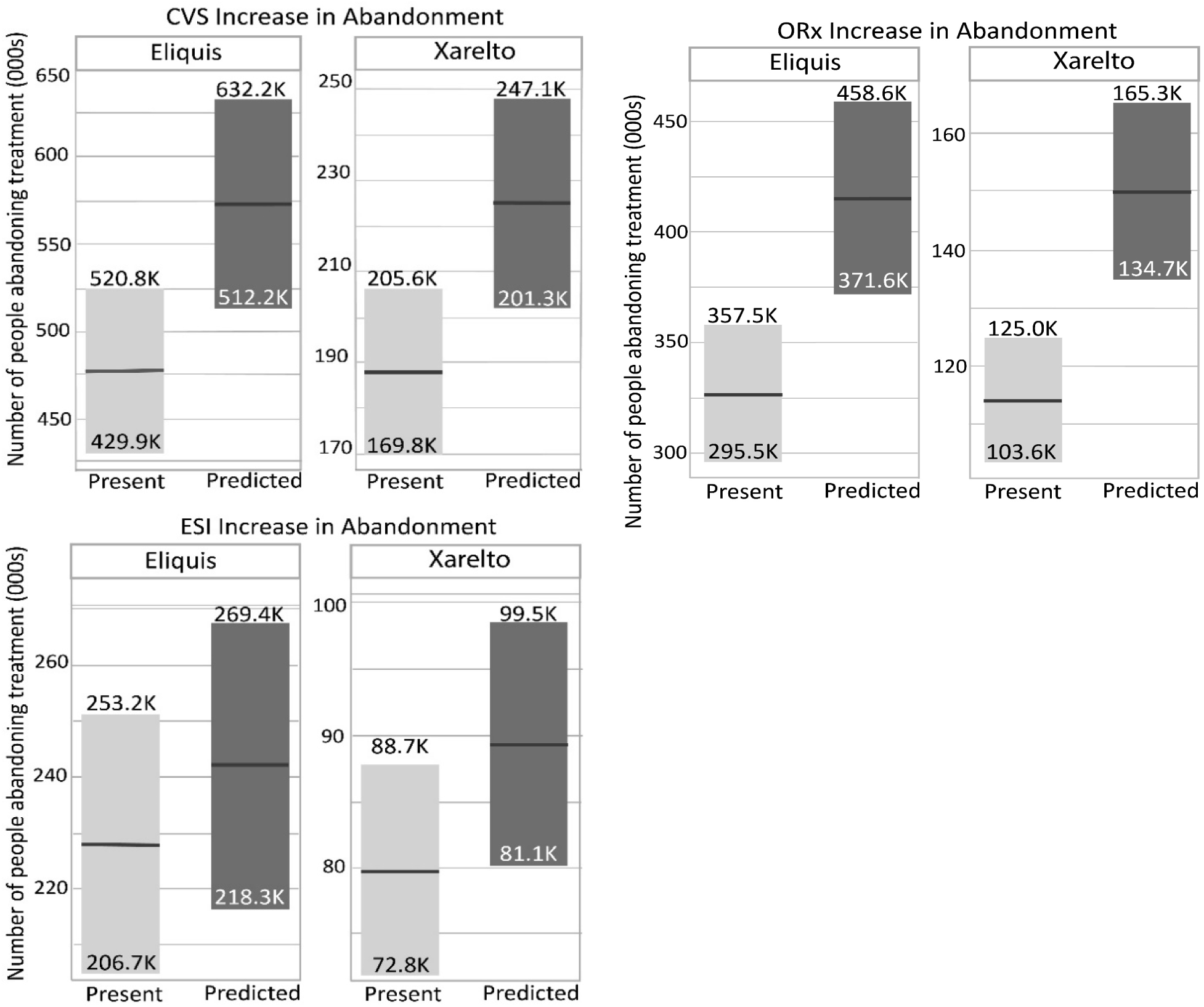
Based on data from IQVIA, regarding the proportion of patients who abandon treatment as copay costs rise,^24^ the likely abandonment rates for Eliquis and Xarelto in 2023 (Present) and if those prescriptions were moved to formulary Tier 6 (Predicted) were calculated. The increased abandonment rates for Eliquis and Xarelto are shown for each of the three largest PBMs, CVS Caremark (CVS), Optum Rx (ORx), and Express Scripts International (ESI), which together manage more than 80% of all prescriptions in the U.S.

Suppose the three largest PBMs shifted Medicare Part D subscriptions for Eliquis or Xarelto to Tier 6, a surrogate for an increase in out-of-pocket cost. In that case, we estimate between 169,000 and 228,000 (Eliquis) or 71,000 to 93,000 (Xarelto), more patients would stop taking their medicine (Table 4) because of their increased financial burden of treatment.

**Table 4.** Increased number of patients who would abandon treatment with Eliquis or Xarelto if moved to formulary Tier 6.

|  |  |  |  |  |
| --- | --- | --- | --- | --- |
|  | **Eliquis** |  | **Xarelto** |  |
|  | **Minimum** | **Maximum** | **Minimum** | **Maximum** |
| CVS | 82,000 | 111,000 | 32,000 | 42,000 |
| ORx | 76,000 | 101,000 | 31,000 | 40,000 |
| ESI | 11,000 | 16,000 | 8,000 | 11,000 |
| **Three Largest PBMs** | 169,000 | 228,000 | 71,000 | 93,000 |
| Abbreviations: CVS, CVS Caremark; ESI, Express Scripts International; ORx, Optum Rx; PBM, pharmacy benefit manager. |  |  |  |  |

We utilized a 45% increased risk of major cardiovascular events and a 30% increased risk of all-cause mortality after abandoning anticoagulants,^21^ to estimate 76,000 to 103,000 more major cardiovascular events for patients who abandoned Eliquis and 32,000 to 42,000 events for patients who abandoned Xarelto. Among those abandoning Eliquis and Xarelto, increases in all-cause mortality are projected at 51,000 to 69,000 and 21,000 to 28,000, respectively.

## DISCUSSION

The current PBM contracting model in the U.S. incentivizes higher-priced and higher-rebated medicines, as PBMs retain a percentage of the rebates collected from biopharmaceutical companies as profit. Additionally, higher-priced medicines are preferred since fees are based on the retail price of the covered drug. It is important to note that fees collected by PBMs from manufacturers, such as assessments for specialty drug dispensing through PBM-owned pharmacies, are often neither disclosed nor shared as savings with sponsors nor patients. As the federal government suppresses retail prices of designated medicines through the IRA, the profit margins for PBMs are expected to decrease, even if manufacturers are forced to provide additional concessions. To recoup lost profits, PBMs may increase out-of-pocket costs for seniors, reduce the sharing of rebate concessions with plan sponsors, increase utilization management, or reduce reimbursements to pharmacies. Our modeling suggests the potential impact of increased cost-sharing requirements on patients’ prescription fill behavior and the risk of morbidity and mortality due to treatment abandonment.

The data analyzed show that the three major PBMs—CVS, ORX, and ESI—do not follow a uniform benefit structure for patient cost-sharing requirements. CVS and ORx place Xarelto and Eliquis on Tier 3, with differing percentages of patients at each copay level within this tier. Currently, the benefit models deployed by ORx and CVS are more favorable to patients than ESI, as they have fewer patients who face higher out-of-pocket costs. Consequently, patients requiring anticoagulant treatment may be better off with CVS and ORx plans. However, this also means that CVS and ORx have more room to increase out-of-pocket costs for seniors in the following years. These nuances in benefit design are critical for the Center for Medicaid and Medicare Services (CMS) to consider during price-setting efforts and follow-up monitoring. Past research demonstrates the impact of formulary exclusions on patients.^19–23^ The analysis presented here suggests shifting out-of-pocket costs onto patients, perhaps as a consequence of the IRA MFP, may have a similar detrimental effect. Policies that move patients to higher out-of-pocket spending will not only strain seniors’ finances but also force some to abandon treatments, leading to more severe health consequences, including increased mortality. CMS must consider these consequences when monitoring health plans and scrutinizing the Medicare Part D benefit designs submitted by PBMs for such potential scenarios.

In this analysis, we used changes in medication tiers as a surrogate to examine increased out-of-pocket costs for seniors. However, focusing solely on medication tiers is inadequate. CMS should instead concentrate on the overall distribution of patient out-of-pocket expenditures for the anticoagulant class, regardless of the medication tier. The three PBMs might not shift patients to higher tiers but could increase out-of-pocket costs within the existing tiers.

As with any modeling analysis, the projections reported here are limited by the data available. The actual prices paid for medications by insurers, as negotiated by PBMs, are not publicly available. In addition, only the total number of prescriptions filled by Medicare Part D enrollees was available. These fills had to be assumed as 90-day fills to roughly estimate the number of people taking the medications as one-fourth of all fills. Data on copay amounts were available only as the proportions of filled prescriptions that fell within a range of copays (varying by increments of $50 or $100) for the current Tiers for Eliquis and Xarelto (Tier 3 and Tier 6). As a result, we could only calculate potential ranges of copay increases that would occur if people’s prescriptions moved from Tier 3 to Tier 6. Similarly, because abandonment rates are dependent on copay amounts,^24^ we could only calculate ranges for the number of people who would abandon treatment with such a formulary shift and the number with increased morbidity and mortality as a result of abandonment.

Future research should evaluate formulary patient out-of-pocket requirements and the distribution of patients across tiers for Xarelto and Eliquis following the implementation of IRA price-setting requirements. Furthermore, CMS should assess each patient’s needs individually, as PBMs may shift out-of-pocket costs to other commonly used generic medications.

## CONCLUSION

The implementation of the IRA will offer specific benefits to seniors, such as lower out-of-pocket costs for seniors who hit the $2,000 cap. However, not all outcomes will be favorable, particularly for patients relying on future cures and affordable access to treatments for chronic diseases. As the IRA exerts pressure on PBM’s profits, it may trigger policy shifts that make it harder for patients currently stable on therapy to afford their medicines. To maintain profit margins, PBMs may shift costs to seniors through higher out-of-pocket requirements for both price-controlled drugs and other medications frequently used to manage chronic conditions. Such policies could have devastating effects on patients who depend on these treatments. CMS and policymakers must closely monitor changes in overall affordability and take preemptive measures to ensure that the most vulnerable citizens are not placed in precarious situations leading to poorer health outcomes. Future research should focus on evaluating the impact of formulary design changes on patient out-of-pocket costs and adherence, especially under the new pricing dynamics introduced by the IRA. By proactively addressing these challenges, we can better safeguard the health and well-being of seniors.

## Data Availability

All data produced in the present study are available upon reasonable request to the authors

